# Should the family have a role in deceased organ donation decision-making? A systematic review of public knowledge and attitudes towards organ procurement policies in Europe

**DOI:** 10.1101/2021.09.13.21263252

**Authors:** Alberto Molina-Pérez, Janet Delgado, Michaela Frunza, Myfanwy Morgan, Gurch Randhawa, Jeantine Reiger-Van de Wijdeven, Silke Schicktanz, Eline Schiks, Sabine Wöhlke, David Rodríguez-Arias

## Abstract

**Goal:** To assess public knowledge and attitudes towards the role of the family in deceased organ donation in Europe.

**Methods:** A systematic search was conducted in CINHAL, MEDLINE, PAIS Index, Scopus, PsycINFO, and Web of Science. These databases were last searched on December 15th, 2017. Eligibility criteria were empirical studies conducted in Europe from 2008 to 2017 and addressing either knowledge or attitudes by the public towards the consent system, including the involvement of the family in the decision-making process, for post-mortem organ retrieval. Each record was screened by two or more independent reviewers in three phases. Data collection from each report was performed by two or more independent reviewers.

**Results:** Of the 1,482 results, 467 reports were assessed in full-text form, and 33 were included in this synthesis. Studies show that a majority of the public support the family’s involvement in organ retrieval decision-making and, in particular, their role as surrogate decision-maker when the deceased has expressed no preference.

**Conclusions:** A common conceptual framework and validated well-designed questionnaires are needed to address the role of the family in future studies. The findings should be considered in the development of Government policy and guidance regarding the role of families in deceased organ donation.

## 1. Introduction

Organ transplantation relies on people’s willingness to donate. The two main models of consent for *post-mortem* organ retrieval are opt-in (explicit consent) and opt-out (presumed consent). While in opt-in systems only those individuals who expressed their willingness to donate are considered as potential organ donors, in opt-out systems all adult individuals are deemed organ donors unless they expressly refused while alive. In addition, families play an important role in organ retrieval decision-making. In most countries, the family is asked to make a decision when the deceased had not, that is, when they failed to express any preference regarding donation, and, in some countries, relatives may even be allowed to overrule (veto) the deceased’s explicit consent (1–3).^1^

The role played by the family in organ procurement decision-making may be more consequential than the model of consent (5). To help increase organ donation rates and respect individual autonomy, scholars have proposed to prevent families from overruling their loved one’s intention to donate (6,7). In 2006, the USA amended the Uniform Anatomical Gift Act to restrict the family’s authority to veto the deceased’s first-person authorization. Recently, several opt-out countries, including Argentina (Law 27.447, 2018), Colombia (Law 1805, 2016), France (Law 2016-41, 2017), and Uruguay (Law 18.968, 2013), went a step further by implementing policies to prevent the family from making any decision at all, even when the deceased failed to express their preference regarding organ donation. Previously, only Austria was known to have a similarly restrictive opt-out policy.

The governance quality of national organ procurement policies can be assessed by using indicators such as the levels of public knowledge and support toward these policies (8). This approach is also applicable to policies regarding the role of the family. On the one hand, public awareness over what relatives can and can’t do in the organ transplant system serves as an indicator of the system’s transparency and publicity, which are necessary means for the citizens’ understanding and meaningful engagement in socially controversial health policy debates (8). Public awareness also affects potential donors and their relatives’ autonomy, and it may influence attitudes such as trust and willingness to donate that are crucial for the success of any transplant system (9,10). For example, an information deficit about the role of the family may create an inner tension in the system, hinder the autonomy of the decision to donate, and amplify moral ambivalence and reluctance towards organ donation (11).

On the other hand, public support for the policy serves as an indicator of the policy’s embodiment of people’s shared values and preferences. Organ procurement policies aim to increase the availability of transplantable organs. However, efficiency should not be pursued at the cost of hampering other ends that are valuable to society, such as respect towards individuals’ autonomy and their posthumous interests, and the autonomy and interests of the family (12). Given that individual autonomy, family preferences, and collective interests may conflict with each other, ideal governance in democracy should weigh and integrate as much as possible the competing values and goods of the pluralistic society it serves. Organ donation policy-making may pursue that objective by striving for policies that can be supported by a majority, while minimally hampering the values of opposing minorities (8).

Assessing the governance quality of consent policies for deceased organ procurement, including the role of the family in the decision-making process, requires measurements of the levels of public awareness and public support toward these policies. In 2009, a survey indicated that only 28% of individuals residing in Europe were aware of the laws governing donation and transplantation in their country, but this study was not specific enough to draw any conclusion regarding individual consent or the relatives’ role in organ donation (13). A systematic review (SR) on public awareness and attitudes towards consent for organ donation was conducted in 2008 which focused on opt-out but did not address the role of the family (14). In 2012, the Welsh government conducted an update review (15) as well as a review of the role of families in organ donation (16). Its former report focused on opt-out too and the latter failed to address public awareness and attitudes.

To fill in this gap of knowledge, we conducted a comprehensive SR addressing all consent systems—including opt-in, opt-out, and mandatory choice,—and the role of the family. The main objective of this SR was to measure the levels of public knowledge and support toward these policies. A partial synthesis of results from this SR, focussing specifically on individual consent, has been published elsewhere (17). In this article, we provide a partial synthesis of results that focuses on knowledge and attitudes towards the role of the family. This is the first systematic review conducted on this topic.

## 2. Methods

We followed a seven-step approach for SR of empirical studies in bioethics, including the MIP model (methodology, issues, participants) to define research questions and inclusion/exclusion criteria (18). We also followed the PRISMA 2020 reporting guideline whenever applicable (19). (PRISMA checklist available as supplementary material).

### 2.1. Eligibility criteria

Eligibility criteria were: quantitative and qualitative empirical studies addressing knowledge and/or attitudes towards the systems of consent for post-mortem organ donation by lay people in continental Europe (box 1).

#### Box 1.

Inclusion criteria and search strategy

| Inclusion criteria |  |
| --- | --- |
| Countries: | Any European country, without language restrictions |
| Date of publication: | Between January 2008 and December 2017 |
| Methods: | Empirical studies, quantitative and/or qualitative |
| Sources: | Peer-reviewed journals, grey literature reports |
| Subjects: | General public (i.e. non-healthcare professionals), including |
|  | students |
| Outcomes: | Knowledge and/or attitudes |
| Issues: | —Consent systems (opt-in, opt-out, mandatory choice)<br>—Ways to express individual preferences (registries, donor cards)<br>—Involvement of the family in the decision-making process |
| Conditions: | Donation or procurement<br>Solid organs (no tissues, blood, eggs, etc.)<br>Post-mortem (no living donation)<br>Transplantation (no research) |
| **Databases:** *Ovid*: MEDLINE; *ProQuest*: PAIS Index, PsycINFO; *Ebsco*: CINHAL Complete; *Scopus*; *Web of Science*: all databases. |  |
| **Websites:** Google, health ministries, patients associations, polling firms. |  |
| **Other searches:** As an ancillary search strategy, we checked eligible studies' references, and we consulted experts in the field for any information on potentially eligible reports. |  |
| **Example of a search string** for Ovid MEDLINE(R) In-Process & Other Non-Indexed Citations and Ovid MEDLINE(R) <1946 to Present (December 15th 2017): |  |
| ((public or donor? or famil* or relative? or parent* or next-of-kin) adj4 (decision? or authoriz* or authoris* or accept* or refus* or agree* or disagree* or veto or willing* or override*)).ti,ab,kw. |  |
| This is just one step in a 136-step search strategy. The full algorithm is available in a supplementary file. |  |

**Databases:** Ovid: MEDLINE; ProQuest: PAIS Index, PsycINFO; Ebsco: CINHAL Complete; Scopus; Web of Science: all databases.

**Websites:** Google, health ministries, patients associations, polling firms.

**Other searches:** As an ancillary searchstrategy, we checked eligible studies’ references, and we consulted experts in the field for any information on potentially eligible reports.

**Example of a search string** for Ovid MEDLINE(R) In-Process & Other Non-Indexed Citations and Ovid MEDLINE(R) <1946 to Present (December 15th 2017):

> ((public or donor? or famil* or relative? or parent* or next-of-kin) adj4 (decision?
> or authoriz* or authoris* or accept* or refus* or agree* or disagree* or veto or willing* or override*)).ti,ab,kw.

This is just one step in a 136-step search strategy. The full algorithm is available in a supplementary file.

### 2.2 search strategy

A scoping search was initially run in the PROSPERO database, the Cochrane Library, MedLine, and Google Scholar for similar or related systematic reviews as well as relevant studies. Then, we developed a 136-steps algorithm in Medline that we adapted following the respective conventions and mesh terms of alternative databases (Box 1). We used a wide range of terms and combinations of terms to capture different expressions of the same ideas, including 74 combinations of terms for “consent”, 57 for “deceased organ donation”, and 69 for “knowledge and attitudes”. In addition, we searched for articles and grey literature by browsing the websites of public and private organisations, hand checking relevant publications, and direct contact with experts in the field. Searches were last conducted on December 15th, 2017. The full search strategy is available in the Supplemental File.

### 2.3. Selection process

A team of nine reviewers worked independently in a three-phase screening process of the records retrieved: by title only, by title & abstract, and by full text. Each record was handed to two different reviewers in each phase. A record was excluded when both reviewers in a given phase deemed it irrelevant (as defined by the inclusion criteria in Box 1). In the third phase, a full text record was eligible when both reviewers deemed it relevant. In cases of disagreement, additional reviewers were consulted and a consensus decision was reached. Three records could not be retrieved and assessed by full text.

### 2.4. Data collection and quality assessment

First, one or more reviewers worked in data collection and in quality assessment for each eligible study. Studies were assessed using three criteria applicable to both qualitative and quantitative research with the aim of identifying those that can be considered as “fatally flawed” (Box 2) (20,21). Then, each study was reviewed and discussed by all authors during an in-person working group meeting and a consensus was reached regarding their final inclusion in the SR. Three reports were rated as “fatally flawed” and excluded for the synthesis.

#### Box 2.

Quality assessment criteria

**Primary criteria applicable to both qualitative and quantitative research**

1. Are the aims and objectives of the research clearly stated?
2. Is the research design clearly specified and appropriate for the aims and objectives of the research?
3. Is there a clear account of the process by which their findings were produced?

→ Based on 1 to 3: How would qualify the quality of this study? Sound; Adequate; Unsure; Poor; Fatally flawed.

**Additional criteria applicable to qualitative research**

1. Methods of data collection used? (Focus groups, interviews, other)
2. Is the data analysis methodology described? What is it?
3. Do the authors report the limitations of their study?
4. Is there enough data to support their results/conclusions?

**Additional criteria applicable to quantitative research**

1. Is the questionnaire available or the questions asked directly cited?
2. Is there a response rate given? What is it?
3. Are power calculations reported?
4. Do the authors report the limitations of their study?

### 2.5. Analysis

We found out that family related data was particularly difficult to interpret due to ambiguity regarding the level of authority given to families in organ donation decision-making, as well as discrepancies between laws and actual practices. Such circumstances made it difficult to categorize and assess public awareness on the role of families. In order to increase analytical clarity, we conducted separate analyses. We thus created two datasets. One includes all data relative to the policies for individual consent and the procedures to express a preference (17). The other includes all data relative to the family. This second dataset is the focus of the present article, which builds from previous conceptual and descriptive analyses (1,12,22).

We developed a conceptual framework to clarify and classify the family’s role into four levels of involvement (1). Then, we clustered the data according to the topics addressed and the varying degrees of precision/vagueness of the information provided. This resulted in a bidimensional tree-like structure of questions and answers (Fig. 1). Questions are displayed vertically, from top to bottom, in increasing levels of precision. Answers are displayed horizontally, from left to right, according to the four incremental levels of family involvement.

**Fig. 1.**
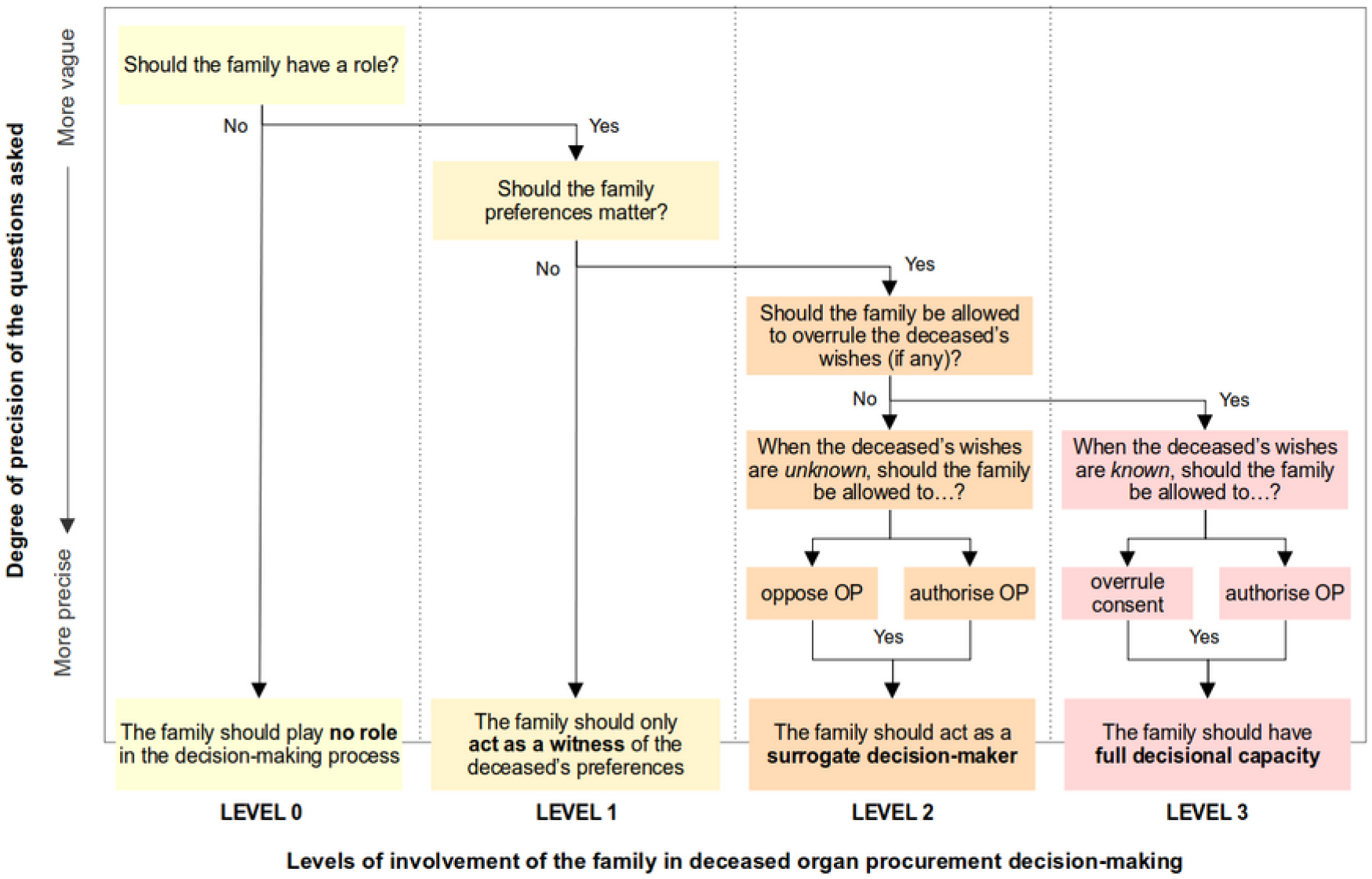
1Tree-like structure of questions and answers according to their degree of precision (vertical) and the level of involvement of the family (horizontal).

Using this scheme, a fresh data extraction was undertaken independently by two reviewers, with any differences discussed with a third reviewer and a consensus reached. If the data from a given study could not answer a question with a sufficient degree of precision, the reviewers went for a less specific question. For instance, if a study claimed that a majority of respondents support the idea that “relatives should decide about organ donation”, this information is not specific enough to respond to the question “Should the family be allowed to overrule the deceased’s wishes (if any)”, but it does answer the less specific question “Should the family preferences matter?”. A few results could not be interpreted unambiguously and were excluded from the dataset.

A statistical metadata analysis was not possible because of the high level of heterogeneity in survey questions and sampling. For this reason, we undertook a descriptive analysis of the quantitative studies following the above scheme (Fig. 1). In addition, we used results from qualitative studies to give quantitative data some perspective.

## 3. Results

The search yielded 1,482 results, with 467 reports assessed in full-text form (Fig. 2). Seventy reports were eventually found eligible. Thirty seven of these studies did not contain any data related to family decision-making and were therefore not included in this synthesis. The remaining 33 studies are composed of 19 articles (Table 1) and 14 grey-literature reports (Table 2).

**Table 1:** Eligible peer reviewed articles

| Ref. | Author, date | Country | Data collection | Field work | Sample (response rate %) | Participants | Sampling / recruitment |
| --- | --- | --- | --- | --- | --- | --- | --- |
| (23) | Bruce, 2013 | Scotland | Questionnaire | 2011 | 200 (~75) | Patients and relatives in emergency department | Convenience |
| (24) | Coppen, 2010 | Netherlands | Questionnaire | 2004, 2007 | 669 (98), 1,094 (80) | Dutch Health Care Consumer Panel | Representative |
| (25) | de Groot, 2015 | Netherlands | Interviews | 2008-2012 | 21 | Relatives of potential deceased organ donors | Purposive |
| (26) | Decker, 2008 | Germany | Telephone interviews | 2005 | 1,000 (n/d) | General public | Representative |
| (27) | Durczynski, 2011 | Poland | Questionnaire | n/d | 170 (n/d) | Small town secondary school students | n/d |
| (28) | Durczynski, 2013 | Poland | Questionnaire | n/d | 100 (n/d) | Male prisoners | n/d |
| (29) | Joshi, 2011 | United Kingdom | Questionnaire | n/d | 439 (84) | University students: South Asian and Whites | Convenience |
| (30) | Katsari, 2015 | Greece | Questionnaire online | 2013-2014 | 1,116 (77) | Students of technical schools | Convenience |
| (31) | Kobus, 2014a | Poland | Questionnaire | n/d | 296 (n/d) | Members of Baptist Church | Diagnostic poll |
| (32) | Kobus, 2014b | Poland | Questionnaire | 2012 | 612 (n/d) | Small and big town residents | Diagnostic poll |
| (33) | Kobus, 2015 | Poland | Questionnaire | 2012 | 395 (n/d) | Rural residents | Diagnostic poll |
| (34) | Kobus, 2016 | Poland | Questionnaire | n/d | 405 (n/d) | Medical and non-medical students | n/d |
| (35) | Lada, 2011 | Croatia | Questionnaire | n/d | 99 (n/d) | Graduate students | n/d |
| (36) | Michalska, 2010 | Poland | Questionnaire | 2008-2009 | 204 (n/d) | Students of religious seminars | Requests sent to the deans |
| (37) | Pawlowicz, 2016 | Poland | Questionnaire | n/d | 1,011 (n/d) | Medical and non-medical university students and high-school students | n/d |
| (38) | Ryckman, 2009 | Netherlands | Questionnaire | n/d | 375 (n/d) | High school students | Participants drawn from another study |
| (39) | Stadlbauer, 2013 | Austria | Questionnaire online | 2012 | 2,025 (24) | Medical and applied sciences students, ICU nurses | Convenience |
| (40) | Webb, 2015 | United Kingdom | Questionnaire | 2013 | 1,549 (n/d) | General public 18+ | Commercial survey panel |
| (41) | Wejda, 2012 | Poland | Questionnaire | n/d | 296 (n/d) | Nuns, priests and students of theological seminaries | n/d |

**Table 2:** Eligible grey literature reports

| Ref. | Author, date | Country | Data collection | Field work | Sample (response rate %) | Participants | Sampling / recruitment | Institution |
| --- | --- | --- | --- | --- | --- | --- | --- | --- |
| (42) | Beaufort Research, 2012 | Wales | Questionnaire | 2012 | 1,006 (n/d) | General public 16+ | Omnibus survey, representative | Welsh Government |
| (43) | Beaufort Research, 2012 | Wales | Focus groups, interviews |  | 52 | Mix of people | Stratified | Welsh Government |
| (44) | Beaufort Research, 2013 | Wales | Questionnaire | 2013 | 1,015 (n/d) | General public 16+ | Omnibus survey, representative | Welsh Government |
| (45) | Beaufort Research, 2014 | Wales | Focus groups, interviews |  | 80 | General public | “free-find” recruitment | Welsh Government |
| (46) | Beaufort Research, 2014 | Wales | Questionnaire | 2013 | 1,022 (n/d) | General public 16+ | Omnibus survey, representative | Welsh Government |
| (47) | Beaufort Research, 2016 | Wales | Questionnaire | 2015<br>2016 | 1,000 (n/d)<br>1,007 (n/d) | General public 16+ | Omnibus survey, representative | Welsh Government |
| (48) | EenVandaag, 2014 | Netherlands | Questionnaire online | 2014 | 18,783 (n/d) | General public | Representative | EenVandaag (television programme) |
| (49) | Ipsos Mori, 2010 | Great Britain | Questionnaire | 2010 | 967 (n/d) | General public 15+ | Omnibus survey | Human Tissue Authority |
| (50) | Ipsos Mori, 2016 | Great Britain | Quantitative | 2016 | 1,001 (n/d) | Adults |  |  |
| (51) | PHA, 2013 | Northern Ireland | Discussion groups, questionnaire | 2013 | 1,012 (n/d) | General public 16+ | Representative | Public Health Agency |
| (52) | Scottish Government, 2017 | Scotland | Qualitative, quantitative | 2016<br>2017 | 824 (n/d) | Individuals and organisations | Online consultation | Scottish Government |
| (53) | Social Market Research, 2015 | Northern Ireland | Questionnaire | 2014 | 1,050 (n/d) | General public 16+ | Representative | Public Health Agency |
| (54) | Welsh Government, 2009 | Wales | Public meetings, written contributions, telephone interviews | 2008<br>2009 | 235, 385 (n/d) | People interested | Survey: representative | Welsh Government |
| (55) | Young, 2017 | Wales | Qualitative, quantitative | 2017 | 1993 (n/d) | Adults | Representative |  |

**Fig. 2.**
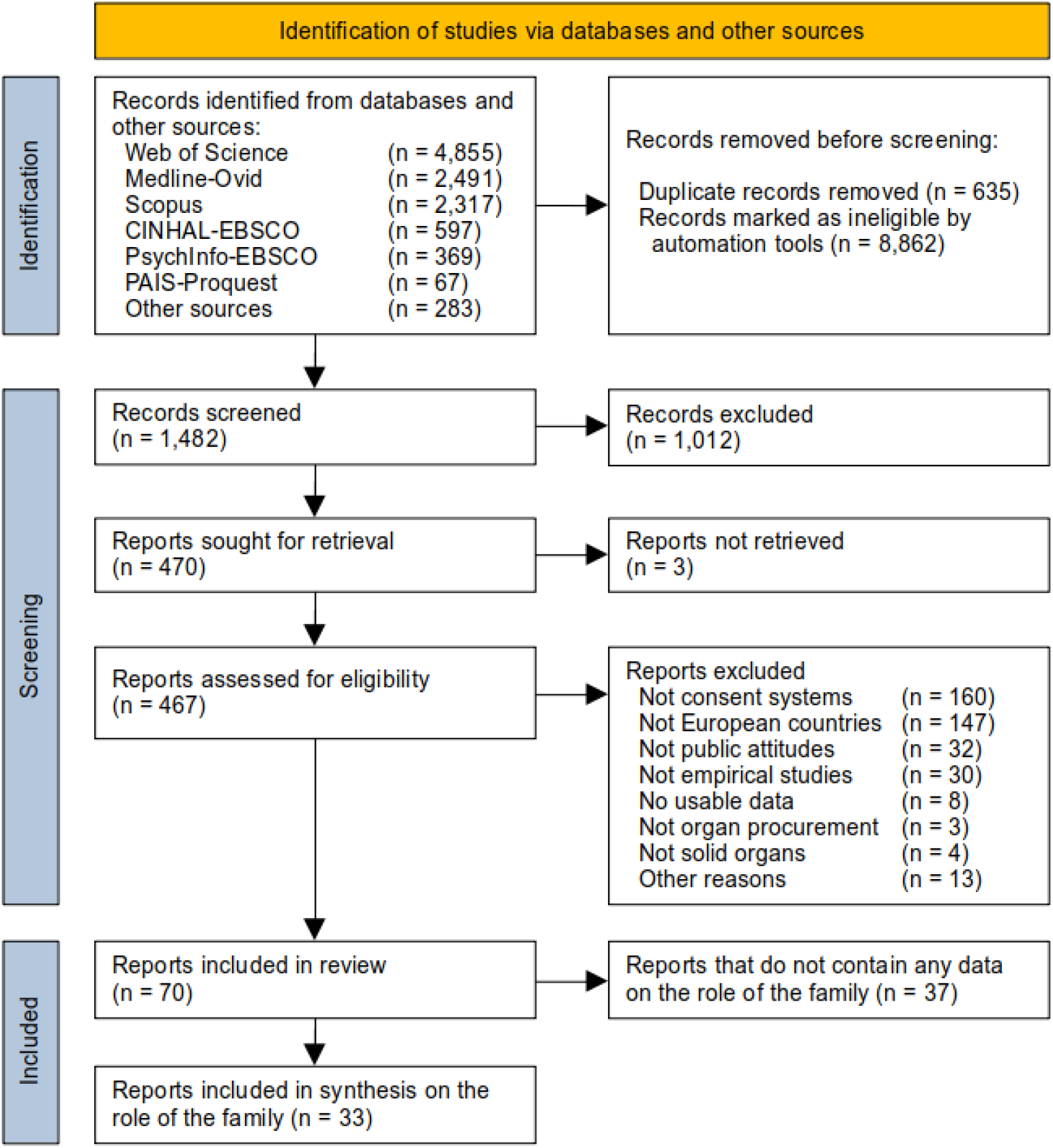
PRISMA 2020 flow diagram (19). Records retrieved from databases and non-database sources were mixed before deduplication and screening. Out of the 70 reports included in review, 37 reports did not contain any data related to the role of the family in decision-making and are therefore not included here. These reports have been included as part of a previous synthesis of results regarding policies for individual consent (17). The remaining 33 reports that do contain relevant data on the role of the family are the focus of the present synthesis.

### 3.1. Public knowledge of the role of the family

We found nine reports with some data on people’s knowledge of the role of the family, including four journal articles (36–38,40), four reports from the Welsh government (43,44,46,54), and one private poll (48). Quantitative results show that, with some exceptions, a minority of the public is aware of the family’s role (Table 3). Qualitative studies in Wales also report people’s ignorance about the family’s capacity to veto a deceased person’s decision to donate (43,54).

**Table 3:** Quantitative results on public knowledge of the role of the family.

| Study | Country | Population | Ratio | Issue (what people know about the role of the family) |
| --- | --- | --- | --- | --- |
| **Can the family override (veto) the deceased's expressed consent?** |  |  |  |  |
| (38) | Netherlands | High school students | 86% | Relatives cannot stop organ donation when someone is registered as an organ donor. |
| (40) | United Kingdom | General population | 48% | A confirmation of consent by the family is required when the potential donor was registered on the organ donation registry |
| (44) | Wales | General population | 45% | The family cannot override the wishes of the deceased |
| (46) | Wales | General population | 46% | The family cannot override the wishes of the deceased |
| **Can the family act as a surrogate decision maker when the deceased failed to express a preference?** |  |  |  |  |
| (40) | United Kingdom | General population | 76% | Relatives are approached to make a decision when the potential donor is not registered |
| (48) | Netherlands | General population | 48% | If the potential donor has not registered a choice on organ donation, relatives have to decide. |
| (36) | Poland | Students | 28% | Family consent is not needed for organ procurement |
| (37) | Poland | Students | 10% | Family consent is not needed for organ procurement |
| (44) | Wales | General population | 51% | If the family is in distress over the decision to donate, clinicians will not proceed with organ donation |
| (46) | Wales | General population | 48% | If the family is in distress over the decision to donate, clinicians will not proceed with organ donation |

### 3.2. Public attitudes towards the role of the family

Results about public attitudes are presented here according to pre-identified four levels of involvement of the family, in decreasing order (from “full authority” to “no role”).

#### Should the family be able to overrule the deceased’s wishes (full authority)?

This question corresponds to the highest level of involvement of the family in organ procurement decision-making (1) (Fig. 1, Level 3). It means that relatives may either authorise the retrieval of organs despite the deceased’s explicit refusal, or oppose the retrieval of organs against the deceased’s explicit consent (also called “family veto”). Thirteen studies addressed this question (24,25,27,29,42,43,45,49,50,52–55). Five addressed it in general terms, eight addressed the specific question of the family’s capacity to veto the deceased’s consent, and none addressed the specific question of the family’s capacity to authorise organ retrieval despite the deceased’s refusal.

Results show that the answer to this question may depend on its wording and the population it is addressed to. When questions are formulated from the perspective of the next-of-kin —i.e. “Taking into account the cited legal conditions, would you [as next of kin of the deceased] approve a situation in which you are not consulted at all?”,—51% to 57% of respondents in the Netherlands answer that they would not approve of not being consulted even if the deceased was registered as a donor (24). Similarly, 53% of respondents in Poland say they “want to be asked for final permission to collect organs from [their] deceased family member” (27). A qualitative study of bereaved relatives in the Netherlands found that, although all families greatly valued the last will of the deceased, a majority felt that “the family has more right to decide about donation” (25).

By contrast, when questions are formulated from the perspective of the donor or in more neutral terms (e.g. *should the families be able to veto their relatives’ expressed consent?*), only 8% to 14% of respondents in the UK support the family’s right to veto and 64% to 88% oppose it (50,52–54). In Wales, 10% to 22% of respondents support the family’s capacity to overrule the deceased’s wishes (42,47,55). This result is consistent with qualitative studies in Wales where participants felt that organ donation was their decision alone and that their family should not be able to overturn it (43,45,54).

When the question is asked to different ethnic groups in the UK, respondents’ support of family veto varies from 9-13% among Whites to 22-27% among Indians, and 46-53% among Pakistanis/Bangladeshis (29). Another study also reports that “black and minority ethnic groups are more equivocal on the question of families overriding a deceased’s wishes”, with 11% strongly objecting to this—against 32% of whites—, and 37% who neither agree nor disagree—against 19% of whites (49).

Results also seem to vary depending on the relationship between the deceased and the decision-maker: respondents were more inclined (by 4% to 7%) to consider that a married next of kin has the right to overrule their deceased spouses’ wishes as compared with parents’ given authority to overrule their deceased offspring’s wishes, and that difference in attitudes was displayed by Indians, Pakistanies/Bangladeshis, and Whites (29).

#### Should the family make a decision when the deceased had not (surrogate role)?

This question corresponds to the second level of involvement of the family (1) (Fig. 1, Level 2). It means that relatives may either authorise or oppose organ retrieval when the deceased had failed to express any preference regarding donation. Six studies addressed this question (24,34,40,49,50,54). In the Netherlands, 66-68% and 80% of the next of kin consider they should be consulted under an explicit consent (opt-in) and a presumed consent (opt-out) system respectively (24). In Poland, to a more neutral question (i.e. “Should the doctor ask the family of the deceased person whether they would accept the removal of organs although the deceased person had not expressed their objection before death?”), 76% of respondents answer that the family should be asked to approve the removal of organs (34). In the UK, 48-50% of respondents thought that the family should decide on behalf of the deceased (49,50,54). Conversely, also in the UK, 30% of respondents answered that “I don’t have the right to make that decision for someone else” (when the deceased is not registered and they had not discussed wishes) (40).

#### Should the family be able to make any decision at all?

This question corresponds to the more basic levels of involvement of the family, in which relatives play no role at all or only act as witnesses of the deceased’s wishes (Fig. 1, levels 0 and 1). Eleven studies asked if the family should make a decision on organ procurement without any specification of the circumstances of that decision (26,28,30–33,35,39–41,52). Affirmative answers suggest that, according to respondents, the family should play a role in the decision-making process and that their own preferences should matter. The ratio of affirmative answers is also extremely variable depending on the study and the population asked, ranging from 8% to 81%. Negative answers are more eloquent, as they suggest that, according to respondents, relatives should *not* make any decision and should therefore either play no role at all (Level 0) or act only as witnesses of the deceased’s wishes by communicating to health professionals the most updated wishes of the deceased, without expressing their own preferences (Level 1). In a study on prisoners in Poland, 65% “answered that only donors could make a decision regarding donation” (28). In all other studies reporting negative answers, the proportion of respondents who thought that the family should not make a decision was much lower: around 20% in Poland (32,33) and the UK (40), around 30% in Croatia (35) and Austria among students (39), and 46% in Austria among transplantation patients (39).

#### Other results

Some studies explored issues not included in the above synthesis. In particular, one study explored the attitudes of patients and relatives in the emergency department with regard to the acceptability of preservation procedures being carried out to maintain organ viability both before and after discussion with relatives (23). Other issues include whether or not registration in the organ donor register without having spoken with relatives is sufficient as an expression of the individual’s wishes (40), whether participants would register an objection if the soft opt-out system was introduced (51), and whether families are capable of making decisions due to their emotional situation (52).

## Discussion

Overall, studies show that a majority of the public supports the family’s involvement in organ retrieval decision-making and, in particular, their role as surrogate decision-maker when the deceased has expressed no preference (a circumstance that may be quite frequent).

We provide two possible interpretations for this result. First, relatives may be perceived (by themselves and by others) as the most reliable authority to protect the posthumous interests of their loved ones. Second, as relatives can be directly affected by the decision, either positively or negatively, some perceive it as legitimate that they have a say.

Although individual consent can be considered the cornerstone of organ donation ethics (56), the notion of relational autonomy may better fit the connection between individual choice and family decisions as guardians of the deceased’s beliefs and values (12). This notion accounts for the fact that people’s autonomy, needs, and interests are shaped by their social relations with others (57,58). Since human beings are socially embedded, their personal decisions also take place in a social context (59–63). Relational autonomy shifts the attention from the protection of individual choices against others’ interference towards the construction of relations which nurture autonomous decisions (64,65).

As mentioned in the introduction, Argentina, Colombia, France, and Uruguay recently changed their opt-out policies to prevent the family intervention in the decision over organ donation. Some may consider such policy shifts as ethically challenging to the extent that opt-out policies may require “widespread and vigorous public education to ensure understanding, along with clear, easy, non-burdensome, and reliable ways for individuals to register dissent” and, “in view of the difficulty of interpreting silence […] organ recovery teams also consult the decedent’s family” (66). Importantly, a previous result of our systematic review is that, in opt-out countries, public awareness of the need to express dissent and public knowledge of the procedures to do so are, overall, lower than in opt-in countries and, in any case, suboptimal (17). By legally preventing the family from objecting, opt-out countries take the risk of not only frustrating bereaved families’ wishes, but also removing organs from non-donor deceased individuals. This being said, to better assess the governance quality of these countries’ policies with regard to the family, improved measures of public awareness and public support of the policy in question are needed.

Regarding the capacity of the family to overrule the deceased’s explicit wishes, results from this review show a complex picture. When the deceased and the next-of-kin hold conflicting views, the question is whose preferences should prevail. The way people answer this question seems to depend on whether respondents see themselves as potential donors or as a deceased’s next-of-kin. In the first case, a vast majority consider that the deceased’s wishes should be respected, regardless of the family. In the second case, respondents are more divided on whether families should have the last say. In addition, answers may vary depending on the nature of the relationship between the potential donors and the decision-maker(s) within the family, as well as depending on the ethnic or cultural background of the deceased and their family. There is some evidence that family-determination plays a greater role in the East Asian principle of autonomy than it does in the West (67). The relative importance of self-determination and family-determination in organ procurement decisions may also vary between European countries, or depending on age, education, religion, and other psychosocial factors. This requires further investigation.

With regard to knowledge, in many cases, only a minority of the public seems to be aware of the role families play in the decision-making process of organ donation. Results on this matter are limited to four countries and fail to cover all aspects of the family’s role. Hence, this also requires further investigation.

Anyway, measuring people’s awareness of the role of the family in organ procurement proves to be a difficult task because of the opacity over what policy is actually in place in a given country, either *de facto* or *de jure*. A major cause of such opacity is the discrepancy between policy and practice that exists in most jurisdictions, with families usually having a greater role in clinical practice than stated or allowed by the law, as well as the variability of practices from one place to the other (1). An additional cause of opacity is the ambiguity or the silence of several national regulations about the relatives’ rights and duties in this context (22).

A major limitation of this systematic review is the heterogeneity of the sources with regard to the methods employed, as well as the ambiguity and vagueness in studies’ questions and reported results. Indeed, we acknowledge that we have faced considerable difficulties in interpreting the data because of a lack of clarity on the situations in which relatives could be asked to intervene and the different roles they could play. A common conceptual framework to categorize the role of families and properly standardised questionnaires would help to better address this topic in future studies.

## Conclusions

The results of this review indicate that a majority of the public supports the family’s involvement in organ retrieval decision-making and, in particular, their role as surrogate decision-maker when the deceased has expressed no preference. However, a theoretical common framework and validated well-designed questionnaires are needed to address the role of the family in future studies. The findings should be considered in the development of Government policy and guidance regarding the role of families in deceased organ donation.

## Data Availability

All data referred to in the manuscript we publicly available before the start of the study. All data sources are cited.

## Acknowledgments

We thank Anja Marie Bornø Jensen for her contribution in the process of data extraction. We also thank Fátima Al Mesri Rodríguez for her assistance in several parts of the process.

## Ethics statement

All data used in this study were publicly available prior to the initiation of the study. No IRB or ethics committee approval was required.

## Data sharing statement

All data sources used in this study are cited in the manuscript. Requests for further information can be sent to the corresponding author.

## Contributorship statement

AMP, JD and DRA designed and conducted the study and wrote the manuscript. MM and SiSchi supervised the methodology and contributed to eligibility assessment, quality assessment, data extraction, and drafting the manuscript. MF, JR, ES, and SW contributed to eligibility assessment, quality assessment, and data extraction. GR contributed to drafting the report. All authors read and approved the final manuscript.

## Footnotes

1 We use the terms family and relatives in a broad sense, meaning those individuals involved in discussing organ procurement with health care professionals and who may have conflicting knowledge and views on the deceased’s preferences, if any. In some countries, such as the United Kingdom and Chile, the substitute decision-maker within the family is determined by law according to a hierarchical list of relatives. More generally, in the context of health care and organ donation, the family can be understood as a ‘collective actor’ with ‘collective autonomy’ (4).

