## Supplementary file for "Should the family have a role in deceased organ donation decision-making? A systematic review of public knowledge and attitudes towards organ procurement policies in Europe"

**Additional file**

Contents

Methods	2

Design and definitions	2

Data sources and searches	2

Study selection	3

Table 1 : Legal systems of consent for deceased OD in Europe	4

Table 2 : inclusion/Exclusion criteria	2

Table 3 : quality assessment criteria	3

Table 4 : data extraction	1

Search strategy	3

Medline (Ovid)	3

Scopus	8

Web of Science (All databases available)	11

CINHAL	14

PSYCINFO–PROQUEST	18

PAIS International – ProQuest	19

Table 5 : studies included in the review	20

### Table **1** : Inclusion/Exclusion criteria

|  | | **Inclusion Criteria**  If AT LEAST ONE of the following: | **Exclusion Criteria**  If ONLY about: |
| --- | --- | --- | --- |
| **Topic** | **PUBLIC** | Lay people, students, families of donors, minorities | - Physicians, nurses, residents, other healthcare professionals |
|  | **CONSENT system** for procurement or donation | **Consent model:** opt-in, opt-out, mandatory choice, required request, organ conscription  **Role of the family:** family involvement in the decision, proxy decision making…  **Way people are allowed to express their preferences:** registries, advance directives (if related to organ procurement), donor cards | - Allocation of organs, organs preservation techniques, medical training, donation rates, …  - Having or not having a donor card, being or not being registered  - ODH (organ donor holder)  - Financial incentives |
|  | **KNOWLEDGE / ATTITUDES** | Knowledge/awareness towards models of consent  Support, rejection, opinion, etc. towards models of consent | - Attitudes towards organ donation or organ transplantation  - Willingness to donate |
|  | **DECEASED** procurement | Deceased donors; donation after brain death; donation after circulatory death | - Living donation |
|  | **SOLID ORGANS** procurement | Heart, kidney, lung, liver, pancreas, intestine | - Tissues: brain, face, hand/extremities, head, whole body, xenotransplantation, skin, eyes, cornea, blood, gamete, embryo, bone marrow, milk, uterus, penis… |
|  | **PROCUREMENT / DONATION** | Organ procurement, harvesting, removal, retrieval, donation | - Organ recipients, immunosuppression, transplantation outcomes, therapies, diagnosis, for research, for education, … |
| **Countries studied** | | any European country^[1]^ | - Russia, Turkey and Israel do not count as European countries. |
| **Study characteristics** | | Empirical peer-reviewed studies, either qualitative or quantitative. Grey literature surveys if the methods are clear and available. | - Editorials, letters, comments, personal opinion, theoretical texts |
| **Language** | | No limit |  |
| **Date of publication** | | January 1st, 2008 to December 15th 2017 |  |

### Table **2** : Quality assessment criteria

**Primary criteria applicable to both qualitative and quantitative research ***

1. Are the aims and objectives of the research clearly stated?
2. Is the research design clearly specified and appropriate for the aims and objectives of the research?
3. Is there a clear account of the process by which their findings were produced?

      → Based on 1 to 3: How would qualify the quality of this study? Sound; Adequate; Unsure; Poor; Fatally flawed

**Additional criteria applicable to qualitative research**

1. Methods of data collection used? (Focus groups, interviews, other)
2. Is the data analysis methodology described? What is it?
3. Do the authors report the limitations of their study?
4. Is there enough data to support their results/conclusions?

**Additional criteria applicable to quantitative research**

1. Is the questionnaire available or the questions asked directly cited?
2. Is there a response rate given? What is it?
3. Are power calculations reported?
4. Do the authors report the limitations of their study?

*	Dixon-Woods M, Cavers D, Agarwal S, et al. Conducting a critical interpretive synthesis of the literature on access to healthcare by vulnerable groups. BMC Med Res Methodol 2006;6:35.

** Morgan M, Kenten C, Deedat S, on behalf of the DonaTE Programme T. Attitudes to deceased organ donation and registration as a donor among minority ethnic groups in North America and the UK: a synthesis of quantitative and qualitative research. Ethnicity & Health 2013;18:367–90. doi:10.1080/13557858.2012.752073.

### Table **3** : Data extraction

| **Questions / topics** | | | **Answers / results** |
| --- | --- | --- | --- |
| **Knowledge or awareness** | **Legislation about OD (in general)** | |  |
|  | **opt-out** | |  |
|  | **opt-in** | |  |
|  | **mandated choice** | |  |
|  | **required request** | |  |
|  | **role of the family** | |  |
|  | **ODC, adv. directives** | |  |
|  | **other** | |  |
|  | **Main results** | |  |
| **Attitudes** | **Changes to the organ donation system** | |  |
|  | **Should relatives be consulted?** | |  |
|  | **Should relatives have right to veto?** | |  |
|  | **Who should take the decision?** | |  |
|  | **Support for opt-out** | |  |
|  | **Support for opt-in** | |  |
|  | **Support for ODC** | |  |
|  | **agreement / disagreement** | **opt-out** |  |
|  |  | **opt-in** |  |
|  |  | **mandated choice** |  |
|  |  | **required request** |  |
|  |  | **role of the family** |  |
|  |  | **ODC, adv. directives** |  |
|  |  | **other** |  |
|  |  | **Main results** |  |
|  | **model preferences** | **opt-out** |  |
|  |  | **opt-in** |  |
|  |  | **mandated choice** |  |
|  |  | **required request** |  |
|  |  | **role of the family** |  |
|  |  | **ODC, adv. directives** |  |
|  |  | **other** |  |
|  |  | **Main results** |  |
|  | **trust / distrust** |  |  |
|  |  | **Main results** |  |
|  | **other** |  |  |
|  |  | **Main results** |  |
| **Relation between knowledge and attitudes** | |  |  |
|  |  | **Main results** |  |
| **Perception of the information received** | |  |  |
|  |  | **Main results** |  |
| **Knowledge or attitudes about consent to interventions to facilitate donation** | |  |  |
|  |  | **Main results** |  |
| **Countries studied** | | **UK** |  |
|  |  | **Poland** |  |
|  |  | **Greece** |  |
|  |  | **Germany** |  |
|  |  | **France** |  |
|  |  | **Sweden** |  |
|  |  | **Austria** |  |
|  |  | **The Netherlands** |  |
|  |  | **Other** |  |

### Search strategy

#### Medline (Ovid)

Database: Ovid MEDLINE(R) In-Process & Other Non-Indexed Citations and Ovid MEDLINE(R) <1946 to Present (December 15th 2017)>

------------------ DECEASED ORGAN PROCUREMENT ----------------------------

1     exp "Tissue and Organ Harvesting"/

2     exp "Tissue and Organ Procurement"/

3     exp Tissue Donors/

4     or/1-3

5     (organ? adj3 (donat* or donor? or nondonor?)).ti,ab,kw.

6     (organ? adj3 (procurement or harvesting)).ti,ab,kw.

7     (organ? adj3 (remov* or retriev*)).ti,ab,kw.

8     or/5-7

9     ((cadaver* or deceased or dead or death) adj3 (donat* or donor? or organ?)).ti,ab,kw.

10     ((posthum* or postmort* or post mortem or post-mortem) adj3 (donat* or donor? or organ?)).ti,ab,kw.

11     ((cadaver* or deceased or dead or death) adj3 (procurement or harvesting)).ti,ab,kw.

12     ((posthum* or postmort* or post mortem or post-mortem) adj3 (procurement or harvesting)).ti,ab,kw.

13     or/9-12

14     ((transplant or transplantation) adj (donor* or organ? or system)).ti,ab,kw.

15     (organ donation adj2 (system or policy or policies)).ti,ab,kw.

16     (organ? adj2 shortage).ti,ab,kw.

17     or/14-16

18     4 or 8 or 13 or 17

------------------ CONSENT ----------------------------

19     exp Informed Consent/

20     exp Presumed Consent/

21     "Dissent and Disputes"/

22     or/19-21

23     Required Request/

24     Advance Directives/

25     Living Wills/

26     Personal Autonomy/

27     or/23-26

28     (consent* or dissent).ti,ab,kw.

29     (opt-in? or opt-out? or optout? or opting-in or opting-out).ti,ab,kw.

30     ("mandated choice" or mandated-choice or "required request" or "required organ donation request").ti,ab,kw.

31     (organ? adj3 conscription).ti,ab,kw.

32     ((public or donor? or famil* or relative? or parent* or next-of-kin) adj4 (decision? or authoriz* or authoris* or accept* or refus* or agree* or disagree* or veto or willing* or override*)).ti,ab,kw.

33     (advance directive? or donor? card?).ti,ab,kw.

34     or/28-33

35     22 or 27 or 34

-------------------------------------------------------------------

36     18 and 35

------------------ REGISTERS ----------------------------

37     ((register? or registry or registries) adj3 (donor? or donate or donation)).ti,ab,kw.

38     ((register? or registry or registries) adj3 (consent or refusal)).ti,ab,kw.

39     ((register? or registry or registries) adj3 (opt-in or opt-out)).ti,ab,kw.

40     ((organ donor or organ refusal) adj database).ti,ab,kw.

41     or/37-40

-------------------------------------------------------------------

42     36 or 41

------------------ ATTITUDES ----------------------------

43     Public opinion/

44     Social perception/

45     or/43-44

46     Morals/

47     Ethics/

48     or/46-47

49     Attitude to Death/

50     Attitude to Health/

51     or/49-50

52     Health Knowledge, Attitudes, Practice/

53     Health Education/

54     Health Behavior/

55     or/52-54

56     (knowledge or aware*).ti,ab,kw.

57     (attitude? or view? or perspective? or opinion? or perception? or belief?).ti,ab,kw.

58     (concern? or trust or fear?).ti,ab,kw.

59     ((endorse* or support or objection or opposition or refusal) adj4 (consent* or opt-in or opt-out or optout or "required request" or "mandated choice" or "organ conscription")).ti,ab,kw.

60     ((agree* or disagree* or endorse* or support or objection or opposition or refusal or readiness) adj4 (donation or procurement or transplantation)).ti,ab,kw.

61     or/56-60

62     45 or 48 or 51 or 55 or 61

------------------ RAW RESULTS ----------------------------

63     42 and 62

------------------ FILTERING: 1ST STAGE ----------------------------

64     limit 63 to yr="2008 - 2017"

65     (editorial or historical-article or historical-biography or comment or letter).pt.

66     64 not 65

67     "Surveys and Questionnaires"/

68     Empirical Research/

69     Qualitative Research/

70     Focus Groups/

71     (study or studies or survey? or empirical or questionnaire? or interview? or panel or focus group? or qualitative approach or respondents).ti,ab,kw.

72     or/67-71

73     45 or 72

74     66 and 73

75     (Canad* or Mexic* or "United States" or USA).ti,kw.

76     (China or Chinese or Hong Kong or India? or Indonesia* or Japan* or Korea? or Malaysia* or Taiwan*).ti,kw.

77     (Argentin* or Brazil* or Chile*).ti,kw.

78     (Australia? or "New Zealand" or "New Zealanders" or Maori?).ti,kw.

79     (Turk* or Israel* or Iran* or Russia? or Saudi Arabia*).ti,kw.

80     (Algeria* or Egypt* or Morocc* or Nigeria* or Ghan* or Senegal* or Tunisia*).ti,kw.

81     (Africa* or America* or Asia*).ti,kw.

82     or/75-81

83     Europe*.ti,kw.

84     (UK or United Kingdom or Engl* or Scot* or Ireland or Irish or Wales or Welsh).ti,kw.

85     (Portug* or Spain or Spanish or Andorra* or France or French or Switzerland or Swiss or Ital* or Malt* or Austria* or German* or Belgi* or Flemish or Luxemb* or Netherlands or Holland or Dutch).ti,kw.

86     (Iceland* or Denmark or Danish or Norw* or Swed* or Finland or Finnish or Eston* or Latvia* or Lithuania*).ti,kw.

87     (Poland or Polish or Czech or Slovakia* or Hungar* or Slovenia*).ti,kw.

88     (Croatia* or Bosnia* or Serb* or Montenegr* or Albania* or Kosov*).ti,kw.

89     (Gree* or Cypr* or Bulgaria* or Romania* or Moldova* or Ukrain* or Belarus*).ti,kw.

90     or/83-89

91     82 and 90

92     82 not 91

93     74 not 92

------------------ FILTERING: 2ND STAGE ----------------------------

94     Animals/

95     Humans/

96     94 not 95

97     93 not 96

98     Tissue donors/

99     Tissue Transplantation/

100     Tissue Preservation/

101     Blood Preservation/

102     Semen Preservation/

103     (tissue? or biological material? or biological sample? or biobank? or cytology).ti,kw.

104     (cell adj (donor? or donat* or transplant* or register? or registry or registries)).ti,ab,kw.

105     or/98-104

106     organ?.ti,ab,kw.

107     (heart or intestine or kidney? or liver or lung? or pancreas or thymus).ti,ab,kw.

108     ((consent? or deceased) adj4 (donat* or procure* or harvest* or remov* or retriev*)).ti,ab,kw

109     or/106-108

110     105 not 109

111     97 not 110

112     exp Living Donors/

113     ((living or alive) adj3 (donor? or donat* or harvest* or procure*)).ti,ab,kw.

114     (living adj (kidney or liver)).ti.

115     or/112-114

116     exp Death/

117     (cadaver* or deceased or dead or death or posthum* or postmortem or post mortem).ti,ab,kw.

118     or/116-117

119     115 not 118

120     111 not 119

------------------ FILTERING: 3RD STAGE ----------------------------

121     (an?esthes* or analges* or drug? or extubat* or intubat* or medication or pharma* or postoperat* or rehabilit*).ti,kw.

122     (cancer or carcinoma or benign* or malign* or hepatitis or sclerosis or alcohol* or depressi* or hypertens* or hypotherm* or virus or infect* or bacteria or vaccin* or HIV or AIDS).ti,kw.

123     (2D or 3D or angiograph* or arthroscop* or echocardio* or endoscop* or imaging or magnetic resonance or radiation or radiotherap* or tomograph*).ti,ab,kw.

124     or/121-123

125     120 not 124

126     (stem-cell? or genetic* or genom* or DNA or RNA or mutation).ti,kw.

127     (sex* consent or condom? or contracept* or fertility or infertility or sterility or pregnan* or procreat* or reproducti* or abort*).ti,kw.

128     (blood* or hemato* or sperm* or semen or gamete? or ovocyte? or oocyte? or egg? or embryo* or bone marrow or milk).ti,kw.

129     (organ? adj2 preservation).ti,kw.

130     (immuno* or HLA* or antigen* or antibod* or protei* or molecul* or metabol*).ti,kw.

131     ((transplant* or clinical) adj outcome?).ti,kw.

132     (risk adj3 (organ? or transplant* or donor? or factor? or patient?)).ti,kw.

133     ((immunization or stroke) adj regist*).ti,kw.

134     (allocation adj (polic* or system*)).ti.

135     or/126-134

136     125 not 135

#### Scopus

(((((((((((TITLE-ABS-KEY((register or registry or registries) w/2 (donor or donate or donation or consent or refusal or opt-in or opt-out))) or (TITLE-ABS-KEY("organ donor database"))) or (((TITLE-ABS-KEY("Tissue and Organ Harvesting" or "Tissue and Organ Procurement" or "Tissue Donors")) or (TITLE-ABS-KEY(organ w/2 (donat* or donor or nondonor))) or (TITLE-ABS-KEY(organ w/2 (procurement or harvesting or remov* or retriev*))) or (TITLE-ABS-KEY((cadaver* or deceased or dead or death or posthum* or postmort* or "post mortem" or post-mortem) w/2 (donat* or donor or organ))) or (TITLE-ABS-KEY((cadaver* or deceased or dead or death or posthum* or postmort* or "post mortem" or post-mortem) w/2 (procurement or harvesting))) or (TITLE-ABS-KEY((transplant or transplantation) w/0 (donor* or organ or system))) or (TITLE-ABS-KEY("organ donation" w/1 (system or policy or policies))) or (TITLE-ABS-KEY(organ w/1 shortage))) and ((TITLE-ABS-KEY("Informed Consent" or "Presumed Consent" or "Dissent and Disputes" or "Required Request" or "Advance Directives" or "Living Wills" or "Personal Autonomy")) or (TITLE-ABS-KEY(consent* or dissent or opt-in or opt-out or optout or opting-in or opting-out)) or (TITLE-ABS-KEY("mandated choice" or mandated-choice or "required request" or "required organ donation request")) or (TITLE-ABS-KEY(organ w/2 conscription)) or (TITLE-ABS-KEY((public or donor or famil* or relative or parent* or next-of-kin) w/3 (decision or authoriz* or authoris* or accept* or refus* or agree* or disagree* or veto or willing* or override*))) or (TITLE-ABS-KEY("advance directive" or "donor card" or "advance directives" or "donor cards"))))) and ((TITLE-ABS-KEY("public opinion" or "social perception" or morals or ethics or "attitude to death" or "attitude to health" or "Health Knowledge, Attitudes, Practice" or "Health Education" or "Health Behavior")) or (TITLE-ABS-KEY(knowledge or aware* or attitude or view or perspective or opinion or perception or belief or concern or trust or fear)) or (TITLE-ABS-KEY((endorse* or support or objection or opposition or refusal) w/3 (consent* or opt-in or opt-out or optout or "required request" or "mandated choice" or "organ conscription"))) or (TITLE-ABS-KEY((agree* or disagree* or endorse* or support or objection or opposition or refusal or readiness) w/3 (donation or procurement or transplantation))))) and not (DOCTYPE(bz or ed or er or le or no or pr))) and (((TITLE-ABS-KEY("Surveys and Questionnaires" or "Empirical Research" or "Qualitative Research" or "Focus Groups")) or (TITLE-ABS-KEY(study or studies or survey or questionnaire or interview or panel or “focus group” or “qualitative approach” or respondents))))) AND NOT (((TITLE(Canad* or Mexic* or "United States" OR USA OR China OR Chinese OR “Hong Kong” OR India* OR Indonesia* OR Japan* OR Korea* OR Malaysia* OR Taiwan* OR Argentin* OR Brazil* OR Chile* OR Australia* OR "New Zealand" OR "New Zealanders" OR Maori* OR Turk* OR Israel* OR Iran* OR Russia* OR “Saudi Arabia” OR Algeria* OR Egypt* OR Morocc* OR Nigeria* OR Ghan* OR Senegal* OR Tunisia* OR Africa* OR America* OR Asia*)) OR (KEY(Canad* or Mexic* or "United States" OR USA OR China OR Chinese OR “Hong Kong” OR India* OR Indonesia* OR Japan* OR Korea* OR Malaysia* OR Taiwan* OR Argentin* OR Brazil* OR Chile* OR Australia* OR "New Zealand" OR "New Zealanders" OR Maori* OR Turk* OR Israel* OR Iran* OR Russia* OR “Saudi Arabia” OR Algeria* OR Egypt* OR Morocc* OR Nigeria* OR Ghan* OR Senegal* OR Tunisia* OR Africa* OR America* OR Asia*))) AND NOT ((TITLE(Europe* OR UK OR “United Kingdom” OR Engl* OR Scot* OR Ireland OR Irish OR Wales OR Welsh OR Portug* OR Spain OR Spanish OR Andorra* OR France OR French OR Switzerland OR Swiss OR Ital* OR Malt* OR Austria* OR German* OR Belgi* OR Flemish OR Luxemb* OR Netherlands OR Holland OR Dutch OR Iceland* OR Denmark OR Danish OR Norw* OR Swed* OR Finland OR Finnish OR Eston* OR Latvia* OR Lithuania* OR Poland OR Polish OR Czech OR Slovakia* OR Hungar* OR Slovenia* OR Poland OR Polish OR Czech OR Slovakia* OR Hungar* OR Slovenia* OR Gree* OR Cypr* OR Bulgaria* OR Romania* OR Moldova* OR Ukrain* OR Belarus*)) OR (KEY(Europe* OR UK OR United Kingdom OR Engl* OR Scot* OR Ireland OR Irish OR Wales OR Welsh OR Portug* OR Spain OR Spanish OR Andorra* OR France OR French OR Switzerland OR Swiss OR Ital* OR Malt* OR Austria* OR German* OR Belgi* OR Flemish OR Luxemb* OR Netherlands OR Holland OR Dutch OR Iceland* OR Denmark OR Danish OR Norw* OR Swed* OR Finland OR Finnish OR Eston* OR Latvia* OR Lithuania* OR Poland OR Polish OR Czech OR Slovakia* OR Hungar* OR Slovenia* OR Poland OR Polish OR Czech OR Slovakia* OR Hungar* OR Slovenia* OR Gree* OR Cypr* OR Bulgaria* OR Romania* OR Moldova* OR Ukrain* OR Belarus*))))) AND NOT (KEY(Animals) AND NOT KEY(Humans))) AND NOT (((KEY(Tissue donors OR tissue transplantation OR tissue preservation OR blood preservation OR semen preservation)) OR (TITLE(tissue OR biological material OR biological samble OR biobank OR cytology) OR KEY(tissue OR biological material OR biological samble OR biobank OR cytology)) OR (TITLE-ABS-KEY(cell w/0 (donor OR donat* OR transplant* OR register OR registry OR registries)))) AND NOT ((TITLE-ABS-KEY(organ OR heart OR intestine OR kidney OR liver OR lung OR pancreas OR thymus)) OR (TITLE-ABS-KEY((consent or deceased) w/3 (donat* or procure* or harvest* or remov* or retriev*)))))) AND NOT (((KEY(living donors)) OR (TITLE-ABS-KEY((living OR alive) w/2 (donor OR donat* OR harvest* OR procure*))) OR (TITLE(living w/0 (kidney OR liver)))) AND NOT ((KEY(death)) OR (TITLE-ABS-KEY(cadaver* OR deceased OR dead OR death OR posthum* OR postmortem OR post mortem))))) AND NOT ((TITLE(an?esthes* OR analges* OR drug OR extubat* OR intubat* OR medication OR pharma* OR postoperat* OR rehabilit* OR cancer OR carcinoma OR benign* OR malign* OR hepatitis OR sclerosis OR alcohol* OR depressi* OR hypertens* OR hypotherm* OR virus OR infect* OR bacteria OR vaccin* OR HIV OR AIDS) OR KEY(an?esthes* OR analges* OR drug OR extubat* OR intubat* OR medication OR pharma* OR postoperat* OR rehabilit* OR cancer OR carcinoma OR benign* OR malign* OR hepatitis OR sclerosis OR alcohol* OR depressi* OR hypertens* OR hypotherm* OR virus OR infect* OR bacteria OR vaccin* OR HIV OR AIDS)) OR (TITLE-ABS-KEY(2D OR 3D OR angiograph* OR arthroscop* OR echocardio* OR endoscop* OR imaging OR magnetic resonance OR radiation OR radiotherap* OR tomograph*)))) AND NOT (((TITLE(stem-cell OR genetic* OR genom* OR DNA OR RNA OR mutation OR sex* consent OR condom OR contracept* OR fertility OR infertility OR sterility OR pregnan* OR procreat* OR reproducti* OR abort* OR blood* OR hemato* OR sperm* OR semen OR gamete OR ovocyte OR oocyte OR egg OR embryo* OR bone marrow OR milk OR immuno* or HLA* OR antigen* OR antibod* OR protei* OR molecul* OR metabol*)) OR (KEY(stem-cell OR genetic* OR genom* OR DNA OR RNA OR mutation OR sex* consent OR condom OR contracept* OR fertility OR infertility OR sterility OR pregnan* OR procreat* OR reproducti* OR abort* OR blood* OR hemato* OR sperm* OR semen OR gamete OR ovocyte OR oocyte OR egg OR embryo* OR bone marrow OR milk OR immuno* or HLA* OR antigen* OR antibod* OR protei* OR molecul* OR metabol*))) OR ((TITLE(organ w/1 preservation)) OR (KEY(organ w/1 preservation))) OR ((TITLE((transplant* OR clinical) w/0 outcome)) OR (KEY((transplant* OR clinical) w/0 outcome))) OR ((TITLE(risk w/2 (organ OR transplant* OR donor OR factor OR patient))) OR (KEY(risk w/2 (organ OR transplant* OR donor OR factor OR patient)))) OR ((TITLE((immunization or stroke) w/0 regist*)) OR (KEY((immunization or stroke) w/0 regist*))) OR ((TITLE(allocation w/0 (polic* OR system))) OR (KEY(allocation w/0 (polic* OR system))))) AND ( LIMIT-TO(PUBYEAR,2017) OR LIMIT-TO(PUBYEAR,2016) OR LIMIT-TO(PUBYEAR,2015) OR LIMIT-TO(PUBYEAR,2014) OR LIMIT-TO(PUBYEAR,2013) OR LIMIT-TO(PUBYEAR,2012) OR LIMIT-TO(PUBYEAR,2011) OR LIMIT-TO(PUBYEAR,2010) OR LIMIT-TO(PUBYEAR,2009) OR LIMIT-TO(PUBYEAR,2008) )

#### Web of Science (All databases available)

DocType=All document types; Language=All languages;

#1 TS=("Tissue and Organ Harvesting" OR "Tissue and Organ Procurement" OR Tissue Donors)

#2 TS=(organ NEAR/3 (donat* or donor or nondonor or procurement or harvesting or removal or retrieval))

#3 TS=((cadaver or deceased or dead or death) NEAR/3 (donat* or donor or organ or procurement or harvesting))

#4 TS=((posthum* or postmort* or "post mortem" or post-mortem) NEAR/3 (donat* or donor or organ or procurement or harvesting))

#5 TS=((transplant or transplantation) NEAR/1 (donor* or organ or system))

#6 TS=("organ donation" NEAR/2 (system or policy or policies))

#7 TS=(organ NEAR/2 shortage)

#8 #7 OR #6 OR #5 OR #4 OR #3 OR #2 OR #1

#9 TS=("Informed Consent" or "Presumed Consent" or "Dissent and Disputes")

#10 TS=("Advance Directives" or "Living Wills" or "Personal Autonomy")

#11 TS= (consent or assent or dissent)

#12 TS=(opt-in$ or opt-out$ or optout$ or opting-in or opting-out)

#13 TS=("mandated choice" or mandated-choice or "required request" or "required organ donation request")

#14 TS=(organ NEAR/3 conscription)

#15 TS=((public or donor or famil* or relatives or parent* or next-of-kin) NEAR/4 (decision or authoriz* or authoris* or accept* or refus* or agree* or disagree* or veto or willing* or override*))

#16 TS=(advance directive or donor card)

#17 #16 OR #15 OR #14 OR #13 OR #12 OR #11 OR #10 OR #9

#18 #17 AND #8

#19 TS=((register or registry or registries) NEAR/3 (donor or donate or donation))

#20 TS=((register or registry or registries) NEAR/3 (consent or refusal))

#21 TS=((register or registry or registries) NEAR/3 (opt-in or opt-out))

#22 TS=(("organ donor" or "organ refusal") NEAR/1 database)

#23 #22 OR #21 OR #20 OR #19

#24 #23 OR #18

#25 TS=(Public opinion or Social Perception or Morals or Ethics or Attitude to Death or Attitude to Health or "Health Knowledge, Attitudes, Practice" or Health Education or Health Behavior)

#26 TS=(knowledge or aware*)

#27 TS=(attitude or view or perspective or opinion or perception or belief)

#28 TS=(concern or trust or fear)

#29 TS=((endorse* or support or objection or opposition or refusal) NEAR/4 (consent* or opt-in or opt-out or optout or "required request" or "mandated choice" or "organ conscription"))

#30 TS=((agree* or disagree* or endorse* or support or objection or opposition or refusal or readiness) NEAR/4 (donation or procurement or transplantation))

#31 #30 OR #29 OR #28 OR #27 OR #26 OR #25

#32 #31 AND #24

#33 PY=(2008 OR 2009 OR 2010 OR 2011 OR 2012 OR 2013 OR 2014 OR 2015 OR 2016 OR 2017)

#34 #32 AND #33

#35 #32 AND #33 Refined by: [excluding] DOCUMENT TYPES: ( LETTER OR EDITORIAL OR PATENT OR NEWS )

#36 TS=("Surveys and Questionnaires" OR "Empirical Research" OR "Qualitative Research" OR " Focus Groups")

#37 TS=(study OR studies OR survey$ OR empirical OR questionnaire$ OR interview$ OR panel OR focus group$ OR qualitative approach OR respondents)

#38 #36 OR #37

#39 #35 AND #38

#40 TI=(Canad* OR Mexic* OR "United States" OR USA OR China OR Chinese OR Hong Kong OR India? OR Indonesia* OR Japan* OR Korea? OR Malaysia* OR Taiwan* OR Argentin* OR Brazil* OR Chile* OR Australia? OR "New Zealand" OR "New Zealanders" OR Maori? OR Turk* OR Israel* OR Iran* OR Russia? OR Saudi Arabia* OR Algeria* OR Egypt* OR Morocc* OR Nigeria* OR Ghan* OR Senegal* OR Tunisia* OR Africa* OR America* OR Asia*)

#41 TS=(Europe* OR UK OR United Kingdom OR Engl* OR Scot* OR Ireland OR Irish OR Wales OR Welsh OR Portug* OR Spain OR Spanish OR Andorra* OR France OR French OR Switzerland OR Swiss OR Ital* OR Malt* OR Austria* OR German* OR Belgi* OR Flemish OR Luxemb* OR Netherlands OR Holland OR Dutch OR Iceland* OR Denmark OR Danish OR Norw* OR Swed* OR Finland OR Finnish OR Eston* OR Latvia* OR Lithuania* OR Poland OR Polish OR Czech OR Slovakia* OR Hungar* OR Slovenia* OR Croatia* OR Bosnia* OR Serb* OR Montenegr* OR Albania* OR Kosov* OR Gree* OR Cypr* OR Bulgaria* OR Romania* OR Moldova* OR Ukrain* OR Belarus*)

#42 #40 NOT #41

#43 #39 NOT #42

#44 TS=(Animals NOT Humans)

#45 #43 NOT #44

#46 TS=("Tissue donors" OR "Tissue Transplantation" OR "Tissue Preservation" OR "Blood Preservation" OR "Semen Preservation")

#47 TI=(tissue$ OR biological material$ OR biological sample$ OR biobank$ OR cytology)

#48 TS=(cell NEAR/1 (donor$ or donat* or transplant* or register$ or registry or registries))

#49 #46 OR #47 OR #48

#50 TS=(organ$ OR heart OR intestine OR kidney$ OR liver OR lung$ OR pancreas OR thymus)

#51 TS=((consent$ OR deceased) NEAR/4 (donat* OR procure* OR harvest* OR remov* OR retriev*))

#52 #50 OR #51

#53 #49 NOT #52

#54 #45 NOT #53

#55 TS=("Living Donors")

#56 TS=((living OR alive) NEAR/3 (donor$ OR donat* OR harvest* OR procure*))

#57 TI=(living NEAR/1 (kidney OR liver))

#58 #57 OR #56 OR #55

#59 TS=(cadaver* or deceased or dead or death or posthum* or postmortem or post mortem)

#60 #58 NOT #59

#61 #54 NOT #60

#62 TI=(an?esthes* or analges* or drug$ or extubat* or intubat* or medication or pharma* or postoperat* or rehabilit* or cancer or carcinoma or benign* or malign* or hepatitis or sclerosis or alcohol* or depressi* or hypertens* or hypotherm* or virus or infect* or bacteria or vaccin* or HIV or AIDS or 2D or 3D or angiograph* or arthroscop* or echocardio* or endoscop* or imaging or magnetic resonance or radiation or radiotherap* or tomograph*)

#63 #61 NOT #62

#64 TI=(stem-cell? or genetic* or genom* or DNA or RNA or mutation or sex* consent or condom? or contracept* or fertility or infertility or sterility or pregnan* or procreat* or reproducti* or abort* or blood* or hemato* or sperm* or semen or gamete? or ovocyte? or oocyte? or egg? or embryo* or bone marrow or milk)

#65 TI=(organ$ NEAR/2 preservation)

#66 TI=(immuno* or HLA* or antigen* or antibod* or protei* or molecul* or metabol*)

#67 TI=((transplant* or clinical) NEAR/1 outcome$)

#68 TI=(risk NEAR/3 (organ$ or transplant* or donor$ or factor$ or patient$))

#69 TI=((immunization or stroke) NEAR/1 regist*)

#70 TI=(allocation NEAR/1 (polic* or system$))

#71 #70 OR #69 OR #68 OR #67 OR #66 OR #65 OR #64

#72 #63 NOT #71

#### CINHAL

S1 (SU "Organ Procurement") OR (SU "Tissue and Organ Harvesting") OR (SU "Transplant Donors")

S2 (organ N3 (donat* or donor or nondonor))

S3 (organ N3 (procurement or harvesting))

S4 (organ N3 (remov* or retriev*))

S5 S2 OR S3 OR S4

S6 ((cadaver* or deceased or dead or death) N3 (donat* or donor or organ))

S7 ((posthum* or postmort* or post mortem or post-mortem) N3 (donat* or donor or organ))

S8 ((cadaver* or deceased or dead or death) N3 (procurement or harvesting))

S9 ((posthum* or postmort* or post mortem or post-mortem) N3 (procurement or harvesting))

S10 S6 OR S7 OR S8 OR S9

S11 (transplant or transplantation) N1 (donor* or organ or system)

S12 (organ donation N2 (system or policy or policies))

S13 organ N2 shortage

S14 S11 OR S12 OR S13

S15 S1 OR S5 OR S10 OR S14

S16 SU "Consent" OR SU "Dissent and Disputes"

S17 SU "Advance Directives"

S18 SU "Autonomy"

S19 SU "Living Wills"

S20 S16 OR S17 OR S18 OR S19

S21 consent* or assent or dissent

S22 opt-in or opt-out or optout or opting-in or opting-out

S23 mandated choice or mandated-choice or required request or required organ donation request or ( organ N3 conscription)

S24 (public or donor or famil* or relative or parent or next-of-kin) N4 (decision or authoriz* or authoris* or accept* or refus* or agree* or disagree* or veto or willing* or override*)

S25 SU "Organ Donor Cards"

S26 advance directive or donor card

S27 S21 OR S22 OR S23 OR S24 OR S25 OR S26

S28 S20 OR S27

S29 S15 AND S28

S30 (register or registry or registries) N3 (donor or donate or donation)

S31 (register or registry or registries) N3 (consent or refusal)

S32 (register or registry or registries) N3 (opt-in or opt-out)

S33 (organ donor or organ refusal) W1 database

S34 S30 OR S31 OR S32 OR S33

S35 S29 OR S34

S36 SU "Public Opinion"

S37 SU "Attitudes"

S38 SU "Morals"

S39 SU "Ethics"

S40 SU "Bioethics"

S41 SU "Health Knowledge"

S42 SU "Attitude to Health"

S43 SU "Attitude to Death"

S44 S36 OR S37 OR S38 OR S39 OR S40 OR S41 OR S42 OR S43

S45 knowledge or aware*

S46 attitude or view or perspective or opinion or perception or belief

S47 concern or trust or fear

S48 (endorse* or support or objection or opposition or refusal) N4 (consent* or opt-in or opt-out or optout or "required request" or "mandated choice" or "organ conscription")

S49 (agree* or disagree* or endorse* or support or objection or opposition or refusal or readiness) N4 (donation or procurement or transplantation)

S50 S45 OR S46 OR S47 OR S48 OR S49

S51 S44 OR S50

S52 S35 AND S51

S53 S35 AND S51 (Date limits)

S54 S53 NOT PT (Anecdote or Biography or Commentary or Editorial or Letter)

S55 (MM "Empirical Research+")

S56 MM "Quantitative Studies+"

S57 MM "Qualitative Studies+"

S58 MM "Interviews+"

S59 MM "Focus Groups"

S60 MM "Systematic Review"

S61 S55 OR S56 OR S57 OR S58 OR S59 OR S60

S62 study or studies or survey or empirical or questionnaire or interview or panel or focus group or qualitative approach or respondents

S63 S61 OR S62

S64 S54 AND S63

S65 TI ( Canad* or Mexic* or "United States" or USA or China or Chinese or Hong Kong or India* or Indonesia* or Japan* or Korea* or Malaysia* or Taiwan* or Argentin* or Brazil* or Chile* or Australia* or "New Zealand" or "New Zealanders" or Maori or Turk* or Israel* or Iran* or Russia* or Saudi Arabia* or Algeria* or Egypt* or Morocc* or Nigeria* or Ghan* or Senegal* or Tunisia* or Africa* or America* or Asia* )

S66 SU ( Canad* or Mexic* or "United States" or USA or China or Chinese or Hong Kong or India* or Indonesia* or Japan* or Korea* or Malaysia* or Taiwan* or Argentin* or Brazil* or Chile* or Australia* or "New Zealand" or "New Zealanders" or Maori or Turk* or Israel* or Iran* or Russia* or Saudi Arabia* or Algeria* or Egypt* or Morocc* or Nigeria* or Ghan* or Senegal* or Tunisia* or Africa* or America* or Asia* )

S67 S65 OR S66

S68 Europe* or UK or United Kingdom or Engl* or Scot* or Ireland or Irish or Wales or Welsh or Portug* or Spain or Spanish or Andorra* or France or French or Switzerland or Swiss or Ital* or Malt* or Austria* or German* or Belgi* or Flemish or Luxemb* or Netherlands or Holland or Dutch or Iceland* or Denmark or Danish or Norw* or Swed* or Finland or Finnish or Eston* or Latvia* or Lithuania* or Poland or Polish or Czech or Slovakia* or Hungar* or Slovenia* or Croatia* or Bosnia* or Serb* or Montenegr* or Albania* or Kosov* or Gree* or Cypr* or Bulgaria* or Romania* or Moldova* or Ukrain* or Belarus*

S69 S67 NOT S68

S70 S64 NOT S69

S71 MM "Tissue Transplantation"

S72 MM "Blood Donors"

S73 TI ( cell W1 (donor or donat* or transplant* or register or registry or registries) )

S74 SU ( cell W1 (donor or donat* or transplant* or register or registry or registries) )

S75 TI ( tissue or biological material or biological sample or biobank or cytology

S76 SU ( tissue or biological material or biological sample or biobank or cytology )

S77 S71 OR S72 OR S73 OR S74 OR S75 OR S76

S78 organ or heart or intestine or kidney or liver or lung or pancreas

S79 (consent or deceased) N4 (donat* or procure* or harvest* or remov* or retriev*)

S80 S78 OR S79

S81 S77 NOT S80

S82 S70 NOT S81

S83 MM "Living Donors"

S84 MM "Transplant donors"

S85 TI ( (living or alive) N3 (donor or donat* or harvest* or procure*) )

S86 S83 OR S84 OR S85

S87 SU "Death" OR MM "Death+"

S88 cadaver* or deceased or dead or death or posthum* or postmortem or post mortem

S89 S87 OR S88

S90 S86 NOT S89

S91 S82 NOT S90

S92 TI ( anesthes* or analges* or drug or extubat* or intubat* or medication or pharma* or postoperat* or rehabilit* or cancer or carcinoma or benign* or malign* or hepatitis or sclerosis or alcohol* or depressi* or hypertens* or hypotherm* or virus or infect* or bacteria or vaccin* or HIV or AIDS or 2D or 3D or angiograph* or arthroscop* or echocardio* or endoscop* or imaging or magnetic resonance or radiation or radiotherap* or tomograph* )

S93 SU ( anesthes* or analges* or drug or extubat* or intubat* or medication or pharma* or postoperat* or rehabilit* or cancer or carcinoma or benign* or malign* or hepatitis or sclerosis or alcohol* or depressi* or hypertens* or hypotherm* or virus or infect* or bacteria or vaccin* or HIV or AIDS or 2D or 3D or angiograph* or arthroscop* or echocardio* or endoscop* or imaging or magnetic resonance or radiation or radiotherap* or tomograph* )

S94 TI ( stem-cell or genetic* or genom* or DNA or RNA or mutation or sex* consent or condom or contracept* or fertility or infertility or sterility or pregnan* or procreat* or reproducti* or abort* or blood* or hemato* or sperm* or semen or gamete or ovocyte or oocyte or egg or embryo* or bone marrow or milk or immuno* or HLA* or antigen* or antibod* or protei* or molecul* or metabol* )

S95 SU ( stem-cell or genetic* or genom* or DNA or RNA or mutation or sex* consent or condom or contracept* or fertility or infertility or sterility or pregnan* or procreat* or reproducti* or abort* or blood* or hemato* or sperm* or semen or gamete or ovocyte or oocyte or egg or embryo* or bone marrow or milk or immuno* or HLA* or antigen* or antibod* or protei* or molecul* or metabol* )

S96 S92 OR S93 OR S94 OR S95

S97 S91 NOT S96

#### PSYCINFO–PROQUEST

((((MJMAINSUBJECT.EXACT.EXPLODE("Tissue Donation") OR TI,AB,SU(organ NEAR/3 (donat* or donor or nondonor or procurement or harvesting or removal or retrieval)) OR TI,AB,SU((cadaver or deceased or dead or death) NEAR/3 (donat* or donor or organ or procurement or harvesting)) OR TI,AB,SU((posthum* or postmort* or "post mortem" or post-mortem) NEAR/3 (donat* or donor or organ or procurement or harvesting)) OR TI,AB,SU((transplant or transplantation) NEAR/1 (donor* or organ or system)) OR TI,AB,SU("organ donation" NEAR/2 (system or policy or policies)) OR TI,AB,SU(organ NEAR/2 shortage)) AND (MJMAINSUBJECT.EXACT.EXPLODE("Informed Consent") OR MJMAINSUBJECT.EXACT.EXPLODE("Advance Directives" OR "Autonomy") OR (TI,AB,SU (consent or assent or dissent)) OR TI,AB,SU(opt-in$ or opt-out$ or optout$ or opting-in or opting-out) OR TI,AB,SU("mandated choice" or mandated-choice or "required request" or "required organ donation request") OR TI,AB,SU(organ NEAR/3 conscription) OR TI,AB,SU((public or donor or famil* or relatives or parent* or next-of-kin) NEAR/4 (decision or authoriz* or authoris* or accept* or refus* or agree* or disagree* or veto or willing* or override*)) OR TI,AB,SU(advance directive or donor card))) OR (TI,AB,SU((register or registry or registries) NEAR/3 (donor or donate or donation)) OR TI,AB,SU((register or registry or registries) NEAR/3 (consent or refusal)) OR TI,AB,SU((register or registry or registries) NEAR/3 (opt-in or opt-out)) OR TI,AB,SU(("organ donor" or "organ refusal") NEAR/1 database))) AND (((MJMAINSUBJECT.EXACT.EXPLODE("Public Opinion") OR MAINSUBJECT.EXACT("Physical Illness (Attitudes Toward)") OR MAINSUBJECT.EXACT("Student Attitudes") OR MAINSUBJECT.EXACT("Consumer Attitudes") OR MAINSUBJECT.EXACT("Implicit Attitudes") OR MAINSUBJECT.EXACT("Adolescent Attitudes") OR MAINSUBJECT.EXACT("Child Attitudes") OR MAINSUBJECT.EXACT("Parental Attitudes") OR MAINSUBJECT.EXACT("Adult Attitudes") OR MAINSUBJECT.EXACT("Male Attitudes") OR MAINSUBJECT.EXACT("Community Attitudes") OR MAINSUBJECT.EXACT("Health Personnel Attitudes") OR MAINSUBJECT.EXACT("Death Attitudes") OR MAINSUBJECT.EXACT("Teacher Attitudes") OR MAINSUBJECT.EXACT("Health Attitudes") OR MAINSUBJECT.EXACT("Female Attitudes")) OR MJMAINSUBJECT.EXACT.EXPLODE("Morality")) OR TI,AB,SU(knowledge or aware*) OR TI,AB,SU(attitude or view or perspective or opinion or perception or belief) OR TI,AB,SU(concern or trust or fear) OR TI,AB,SU((endorse* or support or objection or opposition or refusal) NEAR/4 (consent* or opt-in or opt-out or optout or "required request" or "mandated choice" or "organ conscription")) OR TI,AB,SU((agree* or disagree* or endorse* or support or objection or opposition or refusal or readiness) NEAR/4 (donation or procurement or transplantation)))) AND YR(>=2016)

#### PAIS International – ProQuest

((((MAINSUBJECT.EXACT.EXPLODE("Organ Donation") OR MAINSUBJECT.EXACT("Death") OR MAINSUBJECT.EXACT("Dying")) OR (TI,AB,SU(organ NEAR/3 (donat* or donor or nondonor or procurement or harvesting or removal or retrieval)) OR TI,AB,SU((cadaver or deceased or dead or death) NEAR/3 (donat* or donor or organ or procurement or harvesting)) OR TI,AB,SU((posthum* or postmort* or "post mortem" or post-mortem) NEAR/3 (donat* or donor or organ or procurement or harvesting)) OR TI,AB,SU((transplant or transplantation) NEAR/1 (donor* or organ or system)) OR TI,AB,SU("organ donation" NEAR/2 (system or policy or policies)) OR TI,AB,SU(organ NEAR/2 shortage))) AND (TI,AB,SU (consent or assent or dissent) OR TI,AB,SU(opt-in$ or opt-out$ or optout$ or opting-in or opting-out) OR TI,AB,SU("mandated choice" or mandated-choice or "required request" or "required organ donation request") OR TI,AB,SU(organ NEAR/3 conscription) OR TI,AB,SU((public or donor or famil* or relatives or parent* or next-of-kin) NEAR/4 (decision or authoriz* or authoris* or accept* or refus* or agree* or disagree* or veto or willing* or override*)) OR TI,AB,SU(advance directive or donor card))) OR (TI,AB,SU((register or registry or registries) NEAR/3 (donor or donate or donation)) OR TI,AB,SU((register or registry or registries) NEAR/3 (consent or refusal)) OR TI,AB,SU((register or registry or registries) NEAR/3 (opt-in or opt-out)) OR TI,AB,SU(("organ donor" or "organ refusal") NEAR/1 database))) AND YR(>=2016)

1. Included European countries are the following : Albania, Andorra, Austria, Belarus, Belgium, Bosnia, Bulgaria, Croatia, Cyprus, Czech Republic, Denmark, England, Estonia, Finland, France, Germany, Greece, Hungary, Iceland, Ireland, Italy, Kosovo, Latvia, Lithuania, Luxemburg, Malta, Moldova, Montenegro, the Netherlands, Norway, Poland, Portugal, Romania, Scotland, Serbia, Slovakia, Slovenia, Spain, Sweden, Switzerland, Ukraine, United Kingdom, and Wales. Excluded countries are Russia, Kazakhstan, and Turkey because they belong to Asia, although part of their territory is in continental Europe. Microstates with a population smaller than 40.000 have not been searched : Liechtenstein, Monaco, San Marino, and Vatican City.
